# medExtractR: A medication extraction algorithm for electronic health records using the R programming language

**DOI:** 10.1101/19007286

**Authors:** Hannah L. Weeks, Cole Beck, Elizabeth McNeer, Cosmin A. Bejan, Joshua C. Denny, Leena Choi

## Abstract

**Objective:** We developed medExtractR, a natural language processing system to extract medication dose and timing information from clinical notes. Our system facilitates creation of medication-specific research datasets from electronic health records.

**Materials and Methods:** Written using the R programming language, medExtractR combines lexicon dictionaries and regular expression patterns to identify relevant medication information (‘drug entities’). The system is designed to extract particular medications of interest, rather than all possible medications mentioned in a clinical note. MedExtractR was developed on notes from Vanderbilt University’s Synthetic Derivative, using two medications (tacrolimus and lamotrigine) prescribed with varying complexity, and with a third drug (allopurinol) used for testing generalizability of results. We evaluated medExtractR and compared it to three existing systems: MedEx, MedXN, and CLAMP.

**Results:** On 50 test notes for each development drug and 110 test notes for the additional drug, medExtractR achieved high overall performance (F-measures > 0.95). This exceeded the performance of the three existing systems across all drugs, with the exception of a couple specific entity-level evaluations including dose amount for lamotrigine and allopurinol.

**Discussion:** MedExtractR successfully extracted medication entities for medications of interest. High performance in entity-level extraction tasks provides a strong foundation for developing robust research datasets for pharmacological research. However, its targeted approach provides a narrower scope compared with existing systems.

**Conclusion:** MedExtractR (available as an R package) achieved high performance values in extracting specific medications from clinical text, leading to higher quality research datasets for drug-related studies than some existing general-purpose medication extraction tools.

## INTRODUCTION

Electronic health records (EHRs) are a rich source of data for clinical research when information can be extracted accurately and efficiently. As one major class of EHR information, medication information can be used in many studies from aiding in defining a phenotype to determining drug exposure.[1] Detailed medication information is required to perform pharmacokinetic studies modeling the movement of a drug in the body over the course of time, and may allow determination of patient characteristics affecting drug exposure including genotypes via pharmacogenomic studies.[2]

Information on drug regimens is often stored in an unstructured format, such as free-text clinical notes; hence, natural language processing (NLP) methodologies need to be developed to extract medication information accurately. Dosing information such as the strength or amount of a drug as well as how often it is taken are needed to compute relevant quantities like dose given intake and daily dose. In order to use this information to understand patient response to a given drug and improve treatment, care must be taken to build research datasets of the highest quality possible. This requires careful extraction of medication information from unstructured EHR databases, including validation of each step in this process. Such processes may prove particularly beneficial in cases which rely on real-world data rather than randomized studies, e.g., pharmacokinetic/pharmacodynamic analyses in pediatric populations.[3]

We describe medExtractR, an NLP algorithm we developed using R to extract medication information such as strength, dose amount and frequency from clinical notes. Our system provides a targeted approach to identify medication entities (sometimes called “attributes”) that facilitate computation of clinical dose-related quantities. We also compare medExtractR with three existing NLP systems that can be used for medication extraction:

MedEx, MedXN, and CLAMP (Clinical Language Annotation, Modeling, and Processing).[4-6] Each of these three systems is designed for general-purpose medication information extraction.

## BACKGROUND AND SIGNIFICANCE

### Natural language processing for clinical texts

As EHR databases became more common in large health systems, the need to implement information extraction tasks to harvest data stored in unstructured formats grew. Although research related to EHRs has been growing exponentially, publications about NLP have increased only marginally, suggesting that NLP systems may currently be under-utilized in EHR-based research.[7] A recent literature review identified 71 mentions of NLP systems in papers published between 2006 and 2016, a majority of which were rule-based. Hybrid systems combining rule-based and machine learning methodologies were also a popular approach.[8] Researchers have also investigated the additional benefit of combining multiple existing NLP systems in ensemble methods.[9]

Clinical information extraction encompasses a variety of applications. Many NLP tools have previously been developed for various purposes, including MetaMap[10], cTAKES[11], MedLEE[12], and MedTagger[13] among others.[14-17] A common application is phenotyping disease areas, for example by extracting diagnoses from clinical text.[18-19] Information extraction is also used to improve clinical workflows, for example by detecting adverse events during treatment or improving patient management by identifying care coordination activities following hospital discharge.[20-21] Another area of application, and the focus of this paper, is extracting medication information from clinical text sources to perform drug-related studies.

### Natural language processing for medication extraction

Medication information, including characteristics of a dosing regimen, is an important piece in understanding and improving patient treatment. Medication extraction is a variant of information extraction, or the process of creating structured data from an unstructured format within a text source. In this named entity recognition task, specific words or phrases within a clinical text are identified and labeled with the appropriate entity type, for example “drug name” or “strength.”[22] Most medication extraction systems combine lexicon and rule-based approaches, though some incorporate machine learning techniques.[23-24]

In the context of medication extraction, a major focus of clinical NLP systems is on the identification of drug names alone. While these systems were successful at extracting drug names, they often struggled to extract other drug entities such as strength or frequency with which the drug is taken. For instance, Jagannathan et al. compared four commercial NLP systems and found that the systems were able to achieve above 90% F-measure for drug names but not for strength, route, and frequency, with some getting as low as 48.3% on frequency.[25] The extraction of drug information beyond the medication name itself was the focus of the Third i2b2 Workshop on NLP Challenges for Clinical Records, in which 20 teams from multiple countries participated. Overall F-measure scores for the top ten teams ranged from 0.764 to 0.857, indicating room for improvement when extracting this type of information.[26]

In the past decade or so, NLP systems with a focus on extracting medication entities have emerged. MedEx, developed at Vanderbilt University, combines a semantic tagger and Chart parser to identify drug names and entities. It achieved precision, recall, and F-measure above 90% for drug names, strength, route, and frequency in a set of discharge summaries, with slightly lower performance on other types of clinical notes.[4] MedXN is a rule-based system developed at Mayo Clinic to extract drug information and normalize it to RxNorm concept unique identifiers (RxCUI), particularly for use in medication reconciliation contexts, or ensuring consistent medication records across departments and sources. In a comparison with MedEx, MedXN demonstrated higher performance on a set of clinical notes when extracting drug names and entities.[5] CLAMP uses a pipeline architecture to break its process into several steps, including part-of-speech tagging and section header identification. It has flexible capabilities to identify concepts such as treatments and laboratory tests in addition to identifying drug-related information.[6] These NLP systems have had broader scopes in the sense that they are more general-purpose medication extraction algorithms.

For the purpose of developing research-focused datasets such as for pharmacokinetic, pharmacodynamic, or pharmacogenomic studies, we developed medExtractR, a more targeted NLP system which focuses on a specific medication at a time. This system is written in R, a programming language developed and widely used for statistical analysis. MedExtractR is an R package available for download from the Comprehensive R Archive Network (CRAN).[27]

## METHODS

### Data

Two primary drugs of interest were selected as candidates for developing medExtractR: tacrolimus and lamotrigine. Both share a wide dosing range that is titrated to achieve a clinical effect, making them ideal targets for pharmacokinetic studies. Tacrolimus, an immunosuppressive drug commonly used for transplant patients to prevent organ rejection, tends to be simple in how often it is prescribed, typically involving the same dose given twice a day. Lamotrigine, however, is an antiepileptic medication with much more complex and variable prescribing patterns. Patients are prescribed to take the drug two or three times daily, often with different morning, midday, and evening dosages. A third drug, allopurinol, provided an additional test drug but was not used for developing medExtractR. Allopurinol is a commonly used uric acid-lowering drug to treat gout, and often has a simple prescription pattern with a single dose being given once daily.

For each drug, we identified clinical notes from patients who were taking one of tacrolimus, lamotrigine, or allopurinol. From this collection, we randomly selected notes to form training and test sets. Tacrolimus and lamotrigine each had 60 training notes and 50 test notes, while allopurinol had 110 test notes. Clinical notes were extracted from the Synthetic Derivative (SD), a de-identified copy of Vanderbilt University electronic health records. The process for developing and de-identifying the SD have been previously described.[28]

### Description of the medExtractR system

MedExtractR relies on a combination of lexicon dictionaries and regular expression patterns to identify relevant medication information. A schematic of the system is shown in Figure 1 and examples of its application to clinical note excerpts are illustrated in Figure 2. Our system is designed to extract particular medications of interest, rather than all medications mentioned in a clinical note. Once a drug mention is found within a note, medExtractR searches in a surrounding window for entities including drug name, strength, dose amount, dose, intake time, frequency, and last dose time. The term ‘drug mention’ refers to an appearance of a drug name within a clinical note along with its associated entities. Drug mentions are only returned when either key dosing information (strength, dose amount, or dose) or last dose time is found. Thus, phrases with only a drug name present are not extracted.

**Figure 1.**
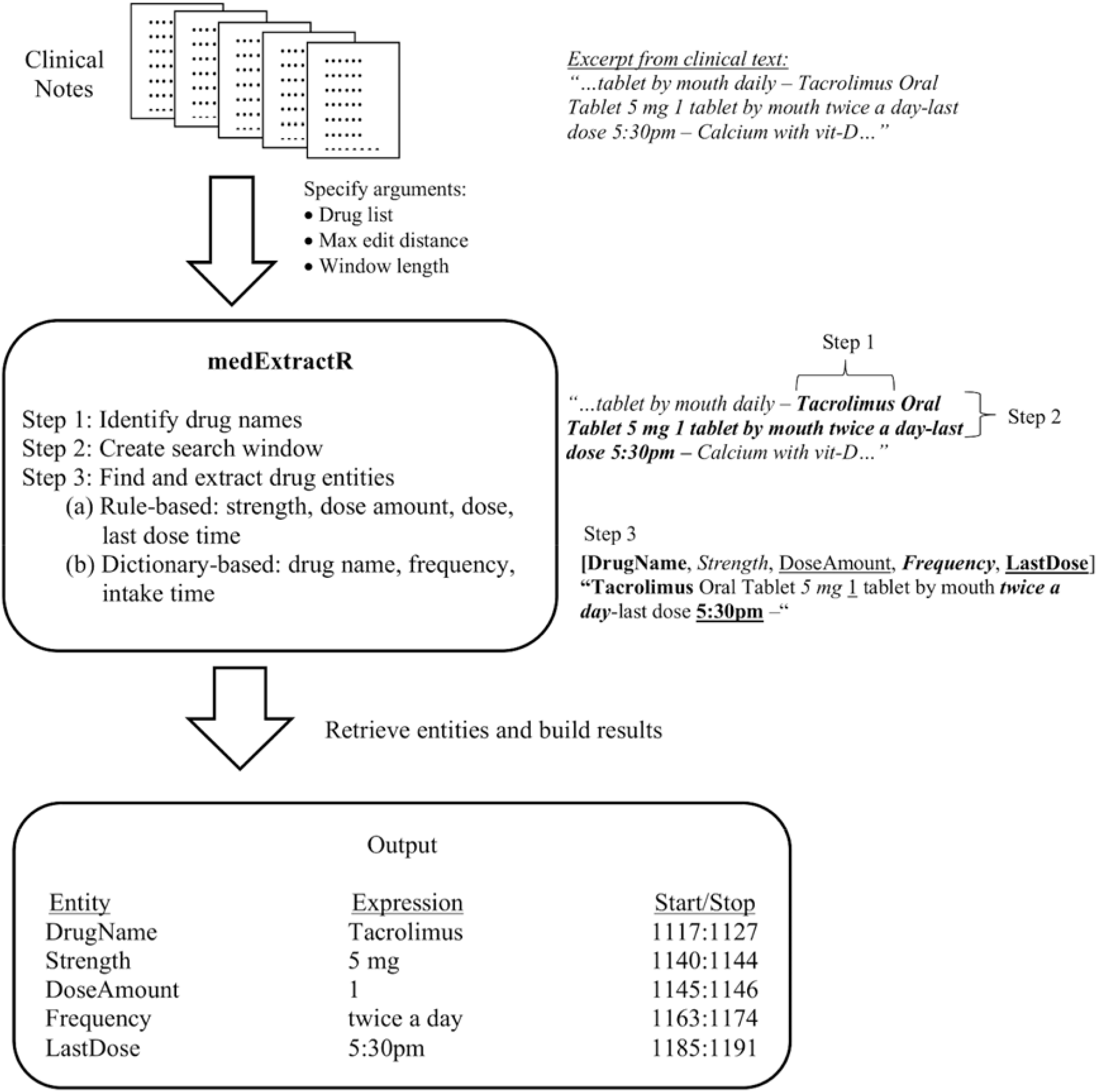
Conceptual representation of the medExtractR system.

**Figure 2.**
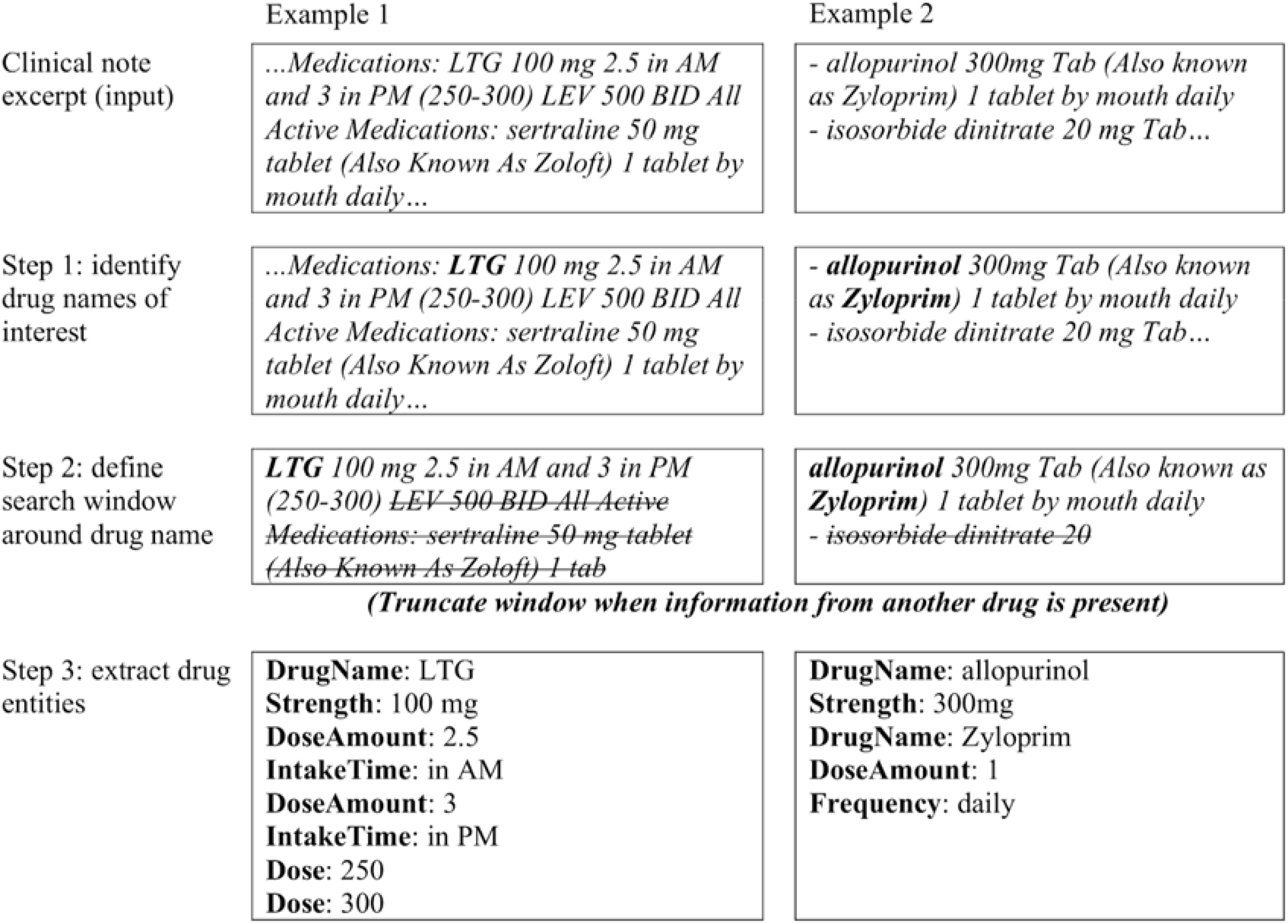
Examples of medExtractR applied to excerpts from clinical notes.

To determine rules for how medExtractR should identify various entities, we manually reviewed the training notes to observe common patterns in how each entity is represented. For some, dictionaries were built based on expressions observed in the training sets while for others, regular expression rules were initially constructed by hand to capture the most commonly observed patterns. We then iteratively improved the dictionaries and rules/regular expressions to maximize performance (F-measure) in the training notes. The following steps outline how medExtractR operates.

#### Step 1: Identify drug names

MedExtractR takes as an argument a vector of names for medications it should extract. Each drug under consideration had its own curated list of names, and was run separately for each list. In contrast to other existing medication extraction NLP systems, medExtractR was developed to extract dosing information for a particular drug of interest to perform medication-specific studies. Thus, each drug list consists of various possible names for a particular drug such as ingredient, brand, and abbreviated names. It then searches within a clinical note to identify all text matching a name from the drug list. For drug names with more than 5 characters, we allow approximate string matching with an edit distance specified by a function argument.

#### Step 2: Create search window

Once a drug mention has been identified within a clinical note, medExtractR then identifies a search window around the mention from which to extract related drug entities. The length of the window (in number of characters) is another function argument and is truncated at the first appearance of an unrelated drug name that is not in the drug list. The ideal window length was chosen to optimize F-measure performance on the training set. This resulted in a window size of 60 characters for tacrolimus and 130 for the more complicated lamotrigine. We used 60 characters for allopurinol since its prescribing patterns are more closely aligned with tacrolimus in simplicity. The list of unrelated drug names was extracted from RxNorm [29] (ingredient ‘IN’ or brand name ‘BN’), from which we removed words which could be confused with regular English words (e.g., ‘today’ or ‘tomorrow’) and supplemented with drug abbreviations observed in the training notes.

#### Step 3: Find and extract drug entities

Within the search window, medExtractR finds and extracts entities of interest, including strength, dose amount, dose, frequency, intake time, dose change, and last dose time.

Descriptions of these entities as well as examples can be seen in Table 1. We require that all drug entities occur within this window with the exception of dose change and last dose time. For dose change, we have a pre-defined list of possible words, e.g., ‘reduce’ or ‘switch,’ observed within the training set. These may indicate a change in dose, and help determine which regimen is current.

**Table 1.** Drug entities identified by medExtractR

| Entity Name | Description | Examples |
| --- | --- | --- |
| Drug Name | Name of the drug for which dose information should be extracted | ‘Prograf’, ‘tacrolimus’, ‘FK-506’, ‘tac’, ‘fk’ |
| Strength | Strength of an individual unit of the medication | ‘5 mg’, ‘100mg’ |
| Dose Amount | Number of units taken at each intake | ‘2’, ‘one’, ‘0.5’ |
| Dose | Total dose given intake | ‘10 mg’, ‘200-200’ |
| Frequency | Number of times per day the drug is taken | ‘once daily’, ‘2x/day’, ‘each day’ |
| Intake Time | Relative time period during the day when a drug is taken | ‘morning’, ‘bedtime’, ‘QPM’, ‘breakfast’ |
| Dose Change | Keyword indicating a change in the dosing regimen | ‘reduce’, ‘increase’, ‘switch’, ‘change’ |
| Last Dose Time | Time expression indicating the time at which the last dose of the drug was taken | ‘8:30 pm’, ‘2030’, ‘830 last night’ |

The remaining entities are identified either using manually curated dictionaries or a combination of regular expressions and rule-based approaches. Last dose time is an optional entity that can occur further from the drug name than dosing information, and as such medExtractR allows extension of the search window for this entity alone via a function argument. Time expressions for last dose can be identified in various formats, including am/pm (“9 pm”), military time (“2100”), or a qualifying expression (“9 last night”), which we identify with regular expressions. In order to be extracted as a last dose time, the search window also must contain a keyword such as “last” or “taken” to reduce false positives.

For both frequency and intake time, we developed dictionaries based on expressions observed within clinical notes and confirmed with physicians when the expressions were ambiguous (e.g., qmon: “one dose per week”; x1: “dosed one time”). Strength, dose amount, and dose primarily rely on regular expressions to identify numeric expressions within the window, e.g., ‘2’ or ‘two’. Simple rules are then used to label the expression as one of these three entities. Strength requires a number followed by a unit that is specified as a function argument, for example “mg.” Examples of rules for dose amount include a number preceding a word like “tablet” or “capsule,” or “take|takes|taking” followed by a number. Dose (i.e., dose given intake) is mathematically equivalent to strength multiplied by dose amount. Dose is often identical in appearance to strength when no dose amount is present within that search window. For one of the development drugs, lamotrigine, we also observed many cases where dose would be written as, e.g., “200-300,” which indicates the patient takes 200 mg in the morning and 300 mg in the evening. In these cases, the presence of the key separation character (“-”) was used to identify both “200” and “300” as dose at different times of day. The code for extracting the drug entities is provided as a function that could be easily customized by the user as needed depending on medication of interest.

### Evaluation

To create a gold standard dataset, we used the brat annotation tool (BRAT) to manually annotate drug entities in clinical notes. [30] We first developed a set of annotation guidelines for each entity by examining how drug information was written within the training notes. Two independent reviewers familiar with the chosen medications independently annotated a set of 20 notes for each of tacrolimus and lamotrigine. We assessed the inter-annotator agreement using Cohen’s kappa separately for each drug. Cases where the two reviewers disagreed were resolved by review from a third expert. The annotation guidelines were then revised to clarify instances that resulted in disagreements.

We annotated a set of 60 training and 50 test clinic notes for each of tacrolimus and lamotrigine. Additionally, we annotated 110 clinic notes for an additional drug, allopurinol, to assess performance on an independent drug not used in medExtractR development. We evaluated medExtractR independently and compared it to three existing clinical NLP systems: MedXN, MedEx, and CLAMP. Performance was assessed using:

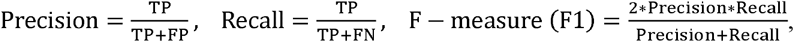

where TP, FP, and FN represent true positives, false positives, and false negatives, respectively. To provide uncertainty in our estimates of precision, recall, and F1, we computed 95% bootstrapped percentile confidence intervals, with 5,000 bootstrap samples at the clinical note level. For independent evaluation of medExtractR, we assessed performance for all drug-entity pairings with the exception of last dose time, which was only evaluated for tacrolimus.

When comparing medExtractR with the existing NLP systems, we standardized the raw output from both the existing systems and medExtractR to ensure compatibility during evaluation. This removed entities not extracted by all systems (e.g., “duration”) and recoded entities having different meaning across systems, for example consolidating medExtractR “dose” and “strength” extractions to all be “strength.” Output generated by the existing NLP systems was manually compared to the gold standard. In cases where another NLP system extracted the same information in the gold standard but represented it differently, we created a validated output field which conformed the extraction to the gold standard representation. For example, in the expression “tacrolimus 1mg 2 capsules twice daily,” MedEx extracts “2 capsules” as dose amount, while the gold standard only has “2.” Another example is that MedXN extracts phrases like “every morning” and “every evening” while medExtractR and the gold standard have just “morning” and “evening.” This manual review ensured that evaluation and comparison across algorithms was fair. The validated entities used in comparing NLP systems were drug name, strength, dose amount, and frequency.

## RESULTS

On 20 notes used for double annotation, the Cohen’s kappa for inter-annotator agreement was 0.970 for tacrolimus and 0.837 for lamotrigine on 84 and 101 annotations, respectively, of labeled medication entities. The lower agreement for lamotrigine was primarily due to titration schedules, or periods of time during which a dose is gradually increased over the course of several days or weeks until it reaches a maintenance dose. In these cases, the reviewers did not initially agree on whether drug entities throughout the entire titrations schedule should be annotated for the gold standard. Annotation guidelines were updated and clarified to resolve any instances of disagreement prior to annotating the training and test sets.

The training sets for tacrolimus and lamotrigine contained 60 notes each, with 105 and 102 drug mentions, respectively. The test sets for tacrolimus, lamotrigine, and allopurinol contained 50, 50, and 110 notes with 88, 76, and 191 drug mentions, respectively. Training set performance for medExtractR was greater than .95 for all drug-entity combinations except lamotrigine-dose and lamotrigine-intake time (Figure 3, Supplementary Table 1). MedExtractR performance remained high on the test set. For most entities, medExtractR achieved precision, recall, and F1 above .90 for tacrolimus and lamotrigine as well as allopurinol, the drug on which it was not trained. Lower performance for intake time may be partially due to relatively low occurrence with only 12 mentions. “Dose” extraction for lamotrigine was highly variable in how it was written in the notes and thus more complicated to extract, resulting in slightly lower performance. Results are not presented for the dose change entity since there were ten or fewer mentions for each of tacrolimus, lamotrigine, and allopurinol. High performance measures (> .95) were also observed for last dose time in tacrolimus notes (Table 2).

**Table 2.** MedExtractR performance for extraction of last dose time on tacrolimus notes.

|  | Training Set<br>(n = 63 extractions) | Test Set<br>(n = 57 extractions) |
| --- | --- | --- |
| Precision | .97 [.92, 1.00] | .98 [.94, 1.00] |
| Recall | .97 [.92, 1.00] | .96 [.91, 1.00] |
| F-measure (F1) | .97 [.92, 1.00] | .97 [.94, 1.00] |

Based on 60 training notes and 50 test notes. Results are presented as “estimate [95% bootstrap confidence interval].”

**Figure 3.**
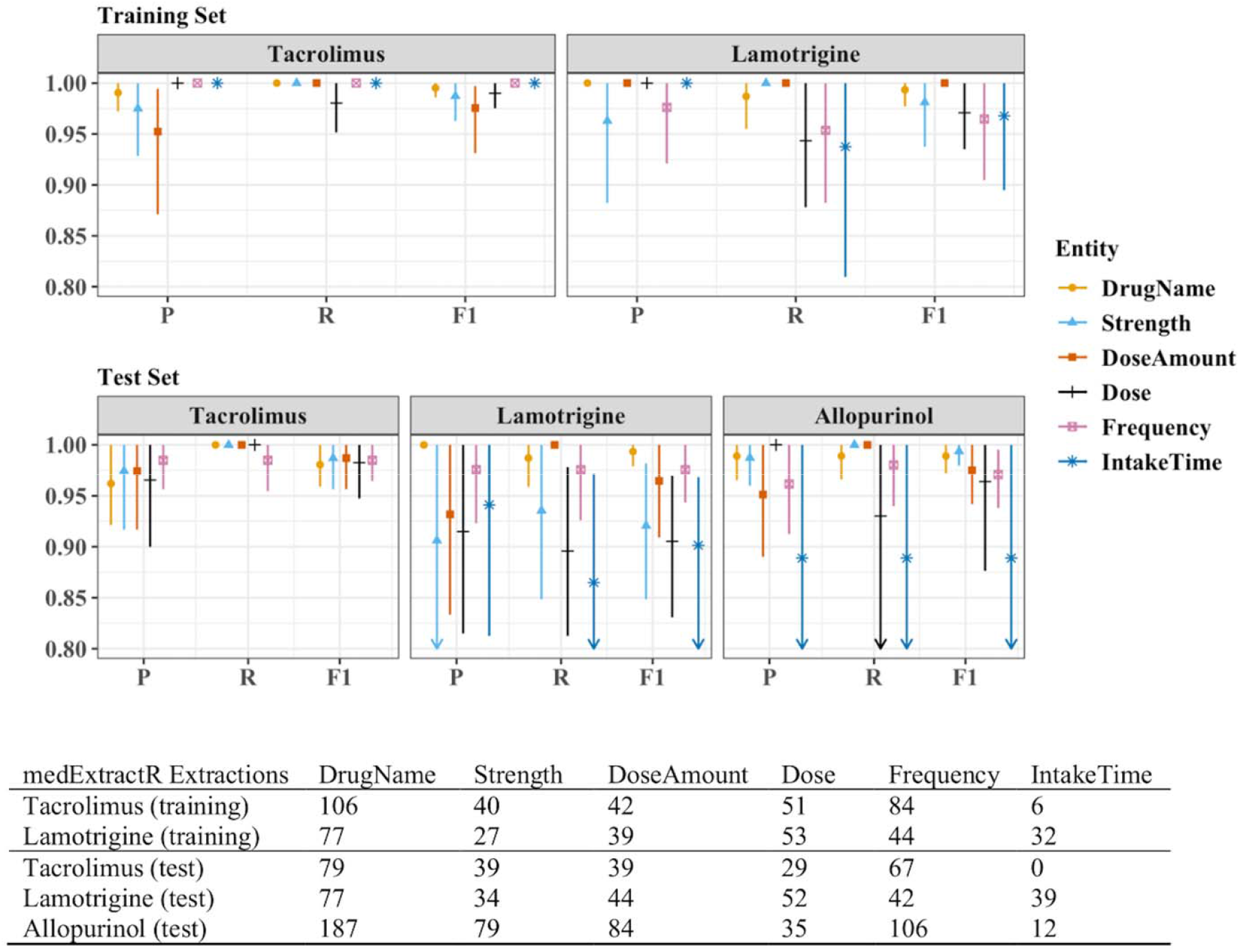
Entity-level precision, recall, and F1 performance measures for medExtractR. The training sets consisted of 60 notes each for tacrolimus and lamotrigine. The test sets consisted of 50 notes each for tacrolimus and lamotrigine, and 110 notes for allopurinol. P, R, and F1 represent precision, recall, and F-measure (F1), respectively. Symbols and lines represent estimates and 95% bootstrapped confidence intervals, respectively. Arrows along the bottom x-axis indicate that either part or all of the confidence interval is below .80. Note that there were no annotations or extractions of intake time for tacrolimus in the test set of clinical notes. Dose change is not shown here since where were ten or fewer mentions for each drug. Numeric results for all entities in this figure and dose change can be found in Supplementary Table 1.

When comparing medExtractR with the other NLP systems, we considered both an overall and entity-level comparison. Here, “overall” means the combined performance aggregating across drug name, strength, dose amount, and frequency entities validated for each NLP system. MedExtractR achieved high overall recall, precision, and F1 (>.95) for tacrolimus and lamotrigine on both the training and test sets as well as for the allopurinol test set. This performance exceeded or matched that of all three existing NLP with respect to F1 for tacrolimus, lamotrigine, and allopurinol (Table 3).

**Table 3.** Precision, recall, and F1 performance measures for medExtractR and three existing NLP systems across standardized and combined drug name, strength, dose amount, and frequency.

| Training Set* Performance |  |  |  |  |  |  |  |  |  |
| --- | --- | --- | --- | --- | --- | --- | --- | --- | --- |
|  | Precision | Tacrolimus<br>Recall | F1 | Precision | Lamotrigine<br>Recall | F1 |  |  |  |
| **medExtractR** | .99 [.98, 1.00] | 1.00 [1.00, 1.00] | .99 [.99, 1.00] | 1.00 [.99, 1.00] | .97 [.94, 1.00] | .98 [.97, 1.00] |  |  |  |
| **MedEx** | .79 [.74, .84] | .74 [.69, .79] | .76 [.72, .81] | .91 [.87, .95] | .73 [.63, .84] | .81 [.74, .87] |  |  |  |
| **MedXN** | .96 [.93, .99] | .90 [.84, .95] | .93 [.89, .96] | .96 [.93, .99] | .76 [.64, .87] | .85 [.76, .92] |  |  |  |
| **CLAMP** | .83 [.77, .88] | .60 [.54, .65] | .70 [.64, .74] | .94 [.90, .97] | .57 [.46, .68] | .71 [.62, .79] |  |  |  |
| Test Set Performance |  |  |  |  |  |  |  |  |  |
|  | Precision | Tacrolimus<br>Recall | F1 | Precision | Lamotrigine<br>Recall | F1 | Precision | Allopurinol<br>Recall | F1 |
| **medExtractR** | .97 [.95, .99] | 1.00 [.99, 1.00] | .98 [.97, 1.00] | .97 [.94, 1.00] | .96 [.91, .99] | .96 [.94, .98] | .96 [.92, .99] | .98 [.96, 1.00] | .97 [.95, .99] |
| **MedEx** | .77 [.71, .84] | .76 [.71, .82] | .77 [.71, .82] | .92 [.87, .96] | .82 [.74, .89] | .87 [.81, .91] | .94 [.90, .97] | .94 [.86, .98] | .94 [.89, .97] |
| **MedXN** | .96 [.92, .98] | .96 [.92, .99] | .96 [.93, .98] | .93 [.89, .96] | .83 [.74, .90] | .88 [.82, .92] | .97 [.94, .99] | .97 [.93, 1.00] | .97 [.94, .99] |
| **CLAMP** | .84 [.78, .91] | .65 [.58, .71] | .73 [.68, .79] | .94 [.90, .97] | .66 [.57, .74] | .78 [.70, .84] | .96 [.93, .99] | .81 [.75, .87] | .88 [.84, .92] |

Results are based on 60 training notes and 50 test notes each for tacrolimus and lamotrigine, and 110 test notes for allopurinol. These are overall results, combining performance across the entities drug name, strength, dose amount, and frequency, which were standardized across systems to ensure comparability. Results are presented as “estimate [95% bootstrap confidence interval].” F1 refers to F-measure.
\*The training set is for medExtractR, and served as another test set for the three existing natural language processing (NLP) systems.

When comparing entity-level extraction, medExtractR often performed as well as or better than the existing NLP systems (Figure 4, Supplementary Table 2). For some drug-entity pairs, including lamotrigine-dose amount and tacrolimus/allopurinol-drug name, medExtractR was outperformed by MedXN due to lower precision. Additionally, low performance on frequency for both tacrolimus and lamotrigine was observed with both MedEx and CLAMP (F1 < 0.8).

**Figure 4.**
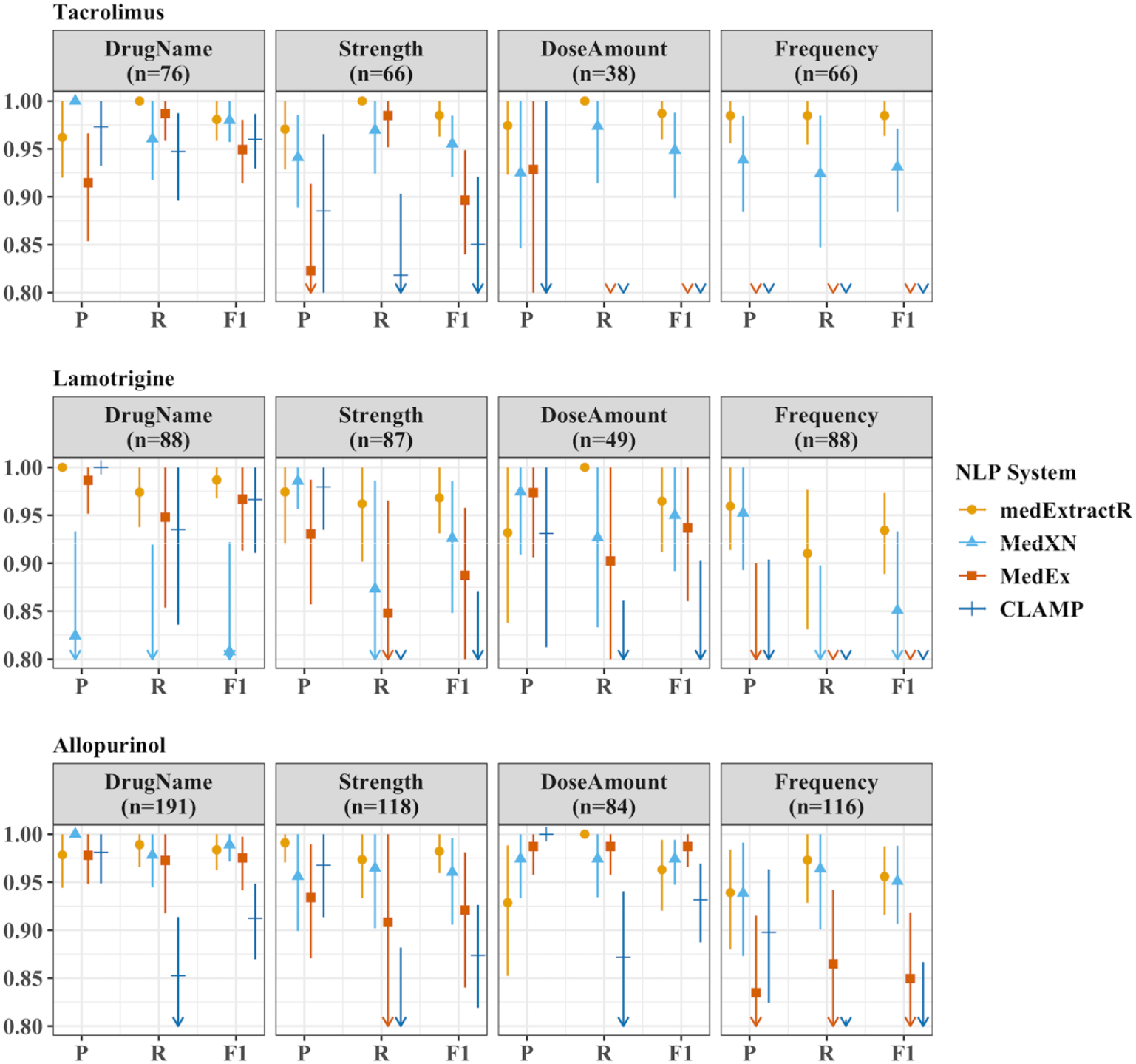
Comparison of medication extraction NLP systems for entity-level precision, recall, and F1 performance measures on test sets. The test sets consisted of 50 notes each for tacrolimus and lamotrigine, and 110 notes for allopurinol. Here, “n” refers to the number of annotations for that drug-entity combination in the gold standard dataset. P, R, and F1 represent precision, recall, and F-measure (F1), respectively. The drug entities presented here reflect a restricted list of entities which have been standardized across all four natural language processing (NLP) systems to ensure comparability. Symbols and lines represent estimates and 95% bootstrapped confidence intervals, respectively. Arrows along the bottom x-axis indicate that either part or all of the confidence interval is below .80. Numeric results for this figure can be found in Supplementary Table 2.

In an error analysis of medExtractR, false negatives often occurred for intake time when exact time expressions were used. We intentionally left out phrases including specific times such as “6 pm” and isolated “am” or “pm.” Including such expressions drastically increased the appearance of false positives in the training set, since they often refer to events which are not of interest, e.g., laboratory times. False negatives also occurred when expressions were not contained in our entity dictionaries, for example the frequency “2x” appeared as a false negative.

False positives occurred when information from a different drug was erroneously extracted. This typically occurred when either the drug was abbreviated but the abbreviation was not included in our supplemental list, or when the name was misspelled. For example, medExtractR returned “75 mg” (strength) and “daily” (frequency) for tacrolimus when these were actually in reference to the drug trazodone, which was misspelled as trazadone in the clinical note.

We also observed errors in the results from the three existing NLP systems we evaluated, many of which were systematic in their occurrence. MedXN failed to identify “LTG” as an abbreviation for lamotrigine, and also did not distinguish between immediate release and extended release formulations. MedEx often failed to identify dose amounts unless they were explicitly indicated with a word like “capsule.” It also consistently extracted “twice a day” as simply “a day.” This latter problem was also shared by CLAMP, although CLAMP did extract the expression “twice a day” under a separate entity called ‘temporal’ which contains many different types of time expressions (e.g., “8 minutes” or “Apr 2014”).

## DISCUSSION

Medication information can be critical data to perform many studies using EHRs. Detailed medication dosing information allows for estimation of drug exposure more accurately via pharmacokinetic studies, or assessment of drug exposure-response relationships via pharmacodynamic studies. Informative models require high quality data, and thus accurate extraction of medication information from EHRs is a pivotal step in advancing research in the medical field. Our results showed high performance for medExtractR both independently and in comparison to three existing NLP systems (MedEx, MedXN, and CLAMP).

MedExtractR operates differently from the existing NLP systems in that it targets specific drugs of interest. In some applications, this may be considered a limitation that it is not optimized for more general medication extraction. In cases such as building research datasets to study particular drugs of interest, however, a system like medExtractR that can achieve higher performance for a narrower scope of medications is desirable. This is analogous to general phenotype exercises, which often employ customized NLP solutions instead of using general-purpose NLP systems (e.g. MetaMap).[1, 10]

MedExtractR not only performed well for the training drugs tacrolimus and lamotrigine, but also demonstrated generalizability with high performance on an additional drug not used in the training data, allopurinol. Different medications are not only subject to variations in prescribing patterns, but also in writing styles between providers. The higher performance seen with medExtractR may partially be due to its ability to better capture such variations through customizable arguments, e.g., the window length parameter. A current limitation of medExtractR is that it was developed for use with oral tablet or capsule medications. A future aim to further expand the usefulness of medExtractR is to determine changes that should be made for extraction of alternative administration routes or dosage forms (e.g., intravenous or oral solution medicine).

We also acknowledge that some existing medication extraction NLP systems, such as MedEx, incorporate an additional “post-processing” step in which individual entities are paired up to provide a dosing regimen given intake. For instance, a note may describe both the patient’s current dose and a new dose being prescribed at that visit. We are currently developing a tool of this nature for both medExtractR and MedXN to construct patient dosing schedules from their entity-level extraction. By separately developing these two procedures (i.e., entity-level dose extraction and pairing of the extracted entities), each of which is optimized and validated, we hope to generate more robust clinical datasets.

## CONCLUSION

In this work we presented medExtractR, a medication extraction algorithm written in R designed to focus on individual drugs for creating research datasets. We demonstrated that medExtractR achieved high performance for three different drugs, and outperformed the existing medication extraction systems MedEx, MedXN, and CLAMP. The ultimate goal of using medExtractR is to develop datasets from EHRs for various medication-related studies requiring more detailed dosing information such as pharmacokinetic/pharmacodynamic or pharmacogenomic analyses. Future work will validate medExtractR’s ability to correctly identify quantities such as dose given intake and daily dose from extracted entities. This information is critical to accurately estimate drug exposure, which can serve as an outcome or an exposure of interest in many drug-related studies.

## Data Availability

Raw data may contain protected health information and are not available to be widely shared. The authors can be contacted for questions or aggregate versions of the data.

## ACKNOWLEDGEMENTS

The software in this paper uses publicly available data courtesy of the U.S. National Library of Medicine (NLM), National Institutes of Health, Department of Health and Human Services; NLM is not responsible for the product and does not endorse or recommend this or any other product.

## AUTHOR CONTRIBUTIONS

As the principal investigator, LC conceived the research and managed the project. HLW developed the medExtractR software, performed analyses, and drafted the manuscript. CB revised the software and implemented as an R package. EM provided quality assurance of data and entity dictionaries. CAB and JD contributed to methods for software development and evaluation. CAB also provided data infrastructure management. All authors provided feedback to improve medExtractR throughout development, and also reviewed and edited the final manuscript.

## REFERENCES

1. Kirby JC, Speltz P, Rasmussen LV, et al. PheKB: a catalog and workflow for creating electronic phenotype algorithms for transportability. J Am Med Inform Assoc 2016;23(6):1046–52.

2. Birdwell KA, Grady B, Choi L, et al. Use of a DNA biobank linked to electronic medical records to characterize pharmacogenomic predictors of tacrolimus dose requirement in kidney transplant recipients. Pharmacogenet Genomics 2012;22(1):32–42.

3. Van SD, Choi L. Real-World Data for Pediatric Pharmacometrics: Can We Upcycle Clinical Data for Research Use? Clin Pharmacol Ther 2019.

4. Xu H, Stenner SP, Doan S, et al. MedEx: a medication information extraction system for clinical narratives. J Am Med Inform Assoc 2010;17(1):19–24.

5. Sohn S, Clark C, Halgrim SR, et al. MedXN: an open source medication extraction and normalization tool for clinical text. J Am Med Inform Assoc 2014;21(5):858–65.

6. Soysal E, Wang J, Jiang M, et al. CLAMP–a toolkit for efficiently building customized clinical natural language processing pipelines. J Am Med Inform Assoc 2017;25(3):331–6.

7. Wang Y, Wang L, Rastegar-Mojarad M, et al. Clinical information extraction applications: a literature review. J Biomed Inform 2018;77:34–49.

8. Kreimeyer K, Foster M, Pandey A, et al. Natural language processing systems for capturing and standardizing unstructured clinical information: a systematic review. J Biomed Inform 2017;73:14–29.

9. Kuo TT, Rao P, Maehara C, et al. Ensembles of nlp tools for data element extraction from clinical notes. AMIA Annu Symp Proc 2016:1880–9.

10. Aronson AR, Lang FM. An overview of MetaMap: historical perspective and recent advances. J Am Med Inform Assoc 2010;17(3):229–36.

11. Savova GK, Masanz JJ, Ogren PV, et al. Mayo clinical Text Analysis and Knowledge Extraction System (cTAKES): architecture, component evaluation and applications. J Am Med Inform Assoc 2010;17(5):507–13.

12. Friedman C, Alderson PO, Austin JH, et al. A general natural-language text processor for clinical radiology. J Am Med Inform Assoc 1994;1(2):161–74.

13. Torii M, Wagholikar K, Liu H. Using machine learning for concept extraction on clinical documents from multiple data sources. J Am Med Inform Assoc 2011;18(5):580–7.

14. Denny JC, Irani PR, Wehbe FH, et al. The KnowledgeMap project: development of a concept-based medical school curriculum database. AMIA Annu Symp Proc 2003;195-9.

15. Goryachev S, Sordo M, Zeng QT. A suite of natural language processing tools developed for the I2B2 project. AMIA Annu Symp Proc 2006:931.

16. Nguyen AN, Lawley MJ, Hansen DP, et al. A simple pipeline application for identifying and negating SNOMED clinical terminology in free text. Proceedings of the Health Informatics Conference (HIC) 2009:188–93.

17. Gold S, Elhadad N, Zhu X, et al. Extracting structured medication event information from discharge summaries. AMIA Annu Symp Proc 2008:237–41.

18. Sada Y, Hou J, Richardson P, et al. Validation of case finding algorithms for hepatocellular cancer from administrative data and electronic health records using natural language processing. Med Care 2016;54(2):e9–e14.

19. Wang M, Cyhaniuk A, Cooper DL, et al. Identification of people with acquired hemophilia in a large electronic health record database. J Blood Med 2017;8:89–97.

20. Rochefort CM, Buckeridge DL, Forster AJ. Accuracy of using automated methods for detecting adverse events from electronic health record data: a research protocol. Implement Sci 2015;10(1):5.

21. Ruud KL, Johnson MG, Liesinger JT, et al. Automated detection of follow-up appointments using text mining of discharge records. Int J Qual Health Care 2010;22(3):229–35.

22. Martin JH, Jurafsky D. Speech and language processing: An introduction to natural language processing, computational linguistics, and speech recognition. Upper Saddle River: Pearson/Prentice Hall; 2009.

23. Patrick J, Li M. High accuracy information extraction of medication information from clinical notes: 2009 i2b2 medication extraction challenge. J Am Med Inform Assoc 2010;17(5):524–7.

24. Li Z, Liu F, Antieau L, et al. Lancet: a high precision medication event extraction system for clinical text. J Am Med Inform Assoc 2010;17(5):563–7.

25. Jagannathan V, Mullett CJ, Arbogast JG, et al. Assessment of commercial NLP engines for medication information extraction from dictated clinical notes. Int J Med Inform 2009;78(4):284–91.

26. Uzuner Ö, Solti I, Cadag E. Extracting medication information from clinical text. J Am Med Inform Assoc 2010;17(5):514–8.

27. R Core Team. R: A Language and Environment for Statistical Computing. Vienna, Austria; 2019. Available from: https://www.R-project.org/.

28. Roden DM, Pulley JM, Basford MA, et al. Development of a large scale de-identified DNA biobank to enable personalized medicine. Clin Pharmacol Ther 2008;84(3):362–9.

29. Nelson SJ, Zeng K, Kilbourne J, et al. Normalized names for clinical drugs: RxNorm at 6 years. J Am Med Inform Assoc. 2011 Jul-Aug;18(4)441–8.

30. Stenetorp P, Pyysalo S, Topić G, et al. BRAT: a web-based tool for NLP-assisted text annotation. Proceedings of the Demonstrations at the 13th Conference of the European Chapter of the Association for Computational Linguistics 2012:102–7.

